# Comparison and analysis of various complementary diagnosis methods for the current situation and problems of COVID-19 diagnosis

**DOI:** 10.1101/2021.01.14.21249620

**Authors:** Hee Jin Huh, Seok Lae Chae, Dong-Min Kim

## Abstract

We evaluated and compared the diagnostic performance of fluorescence immunoassay (FIA) and two types of serological diagnostic tests: enzyme-linked immunosorbent assay (ELISA) and immunochromatographic assay (ICA) for detection of SARS-CoV-2 antigen and antibody to diagnose COVID-19 infections. This study is aimed to analyze and compare the current status and problems of COVID-19 diagnosis and various alternative diagnostic methods that are viable. The enrolled subjects in our study population were tested with real-time polymerase chain reaction (RT-PCR). ELISA and immunochromatographic diagnostic kit were used to diagnose 362 positive and 3010 negative SARS-CoV-2 specimens, and antigen fluorescence immunoassay kit was used on 62 positive and 70 negative SARS-CoV-2 RT-PCR confirmed samples for diagnosis. As a result, categorizing by the patient symptom onset days, PCL COVID19 Total Ab EIA (ELISA) showed the sensitivity of 93.4% from 15 to 21 days, 94.2% from over 22 days, and the specificity of 99.97%. PCL COVID19 IgG/IgM Rapid Gold (ICA) had a sensitivity of 86.9%, 97.4%, and the specificity of 98.14% respectively. PCL COVID19 Ag Rapid FIA sensitivity was 93.8% from 0 to 7 days, 71.4% from 8 to 12 days and specificity was 98.57%. In conclusion, COVID-19 Ab ELISA and ICA, and COVID-19 Ag FIA are all complementary and applicable diagnostic methods to resolve the current problems of COVID-19 diagnosis.

## Introduction

With the first identification of severe acute respiratory syndrome coronavirus 2 (SARS-CoV-2) in December 2019 from Wuhan, Hubei Province, China, coronavirus disease 2019 (COVID-19) was soon after declared as a pandemic in March 2020 by World Health Organization (WHO)^1^. The virus has not ceased to spread until now, resulting in tremendous public health challenges across the world with over 61 million confirmed cases reported worldwide as of 30 November 2020^2, 3^.

COVID-19 diagnostic test samples are obtained from either a patient’s blood or naso-oropharyngeal specimen. Molecular diagnostic tests, such as next generation sequencing or real-time polymerase chain reaction (RT-PCR), have been recommended to detect SARS-CoV-2 from the upper and lower respiratory tract swab samples for COVID-19 diagnosis^3-7^. However, this method requires at least for 6 or more hours, to accurately process and determine the COVID-19 infection of a patient^8^. In many cases where steep COVID-19 surges occur, performing the molecular tests in a timely manner to close the gap between the high number of patients and the limited number of laboratories has been a critical hinderance from devising of efficient containment strategies for COVID-19 in many countries^9^.

In order to solve this problem, serological tests such as enzyme-linked immunosorbent assay (ELISA) and immunochromatography have been developed to detect antibodies and antigens of a virus. These methods now have become commercialized and can be easily accessed. This allows to play a crucial role in overcoming the limitation of molecular diagnostic test as alternative assays with the advantages such as ability to test large numbers of samples more rapidly with less cost and perform an epidemiologic tracing in a short period of time^10, 11^. Proper isolation and timely detection through a rapid diagnosis of COVID-19 is critical to prevent the spreading of infection among the patients with recurrence of COVID-19, and to decrease the associated prevalence and mortality rates due to unforeseen waves of the pandemic^8, 12^. Therefore, we evaluated and compared the diagnostic performance of two types of serological diagnostic test ELISA and ICA for antibody detection and FIA for antigen detection of SARS-CoV-2.

## Methods

### Study Design for serological test

A total of 50 patients diagnosed as COVID-19 positive were enrolled in this study. All enrolled patients were confirmed to be infected with SARS-CoV-2 by the RT-PCR assays using nasopharyngeal swabs. In the case of negative samples, 1072 residual samples were collected from 1072 healthy people assessed by physical examination during early November of 2019, before the pandemic of COVID-19, and the others collected from healthy donors without any COVID-19 symptoms. A total of 3,372 human blood samples (serum), 362 COVID-19 positive samples and 3,010 negative samples were used for this study. Regarding anticoagulant, 3372 serum samples were separated with gel without any anticoagulants. Positive and negative blood samples for COVID-19 were used for the performance evaluation of the PCL COVID-19 Total Ab EIA and the PCL COVID19 IgG/IgM Rapid Gold at DUMC under respective IRB approval (DUIH 2020-03-025-003 and 110757-202003-HR-02-02). The performance evaluation was conducted retrospectively by comparing the information including test results of the RT-PCR COVID-19.

### Study Design for Antigen test

In case of antigen test, we used nasopharyngeal swab samples in viral transport medium (VTM) and the extraction buffer (the PCL COVID19 Ag Rapid FIA kit). A total of 132 nasopharyngeal swab samples stored in VTM, including 62 positives and 70 negatives for the COVID-19, were used for the retrospective performance evaluation of the PCL COVID19 Ag Rapid FIA at 2 institutions under respective IRB approval (S-IRB-2020-004-04-08 and IRB-2020-01-027). Nasopharyngeal swab samples were collected from individuals diagnosed as positive or negative by RT-PCR testing (PowerChekTM2019-nCoV Real-time PCR Kit, Kogenebiotech, Seoul, Korea).

### RT-PCR confirmation assay

Nasopharyngeal swabs were obtained from each patient for RT-PCR, which was performed using the CFX96™ Real-time PCR detection system (Bio-Rad Laboratories, Hercules, CA) with the Allplex™ 2019-nCoV Assay kit (Seegene Inc., Seoul, Korea) according to the manufacturer’s instructions. The assay is designed to detect RdRP and N genes specific for SARS-CoV-2, and E gene for all of *Sarbecovirus* including SARS-CoV-2.

### Serologic test for detection of antibodies against SARS-CoV-2

#### Lateral flow immunochromatographic assay (LFA)

To evaluate SARS-CoV-2 specific IgM and IgG antibodies, the LFA based rapid test (PCL COVID19 IgG/IgM Rapid Gold) was performed following the manufacturer’s instructions. Each sample was tested by one test and readout (positive/negative) interpreted by two operators in parallel.

#### Enzyme-linked immunosorbent assay (ELISA)

To evaluate total antibody response, the ELISA (PCL COVID19 Total Ab EIA test, PCL Inc., Seoul, Korea) was performed following the manufacturer’s instructions. All tests were performed in duplicate, according to the manufacturer’s instructions and Absorbance/cut-off values ≥ 1.0 were considered positive.

#### Fluorescence Immunochromatographic Assay (FIA)

To detect the SARS-CoV-2 virus infection, the rapid FIA kit (PCL COVID19 Ag Rapid FIA), which is used with the PCLOK EZ analyzer for the qualitative detection from SARS-CoV-2 in nasopharyngeal specimens from individuals who are suspected of COVID-19 by their healthcare provider, was performed following the manufacturer’s instrument.

### Statistical analysis

The sensitivity, specificity, positive predictive value (PPV), and negative predictive value (NPV) were calculated, along with the 95% confidence interval (CI). A diagnostic value test of the ELISA and rapid antibody method was performed with MedCalc software, version 19.5.1 (Mariakerke, Belgium).

## Results

### Population of study sample

The overall contents of the study population are described (Table 1). For the Ab test, 362 blood samples were collected from 50 patients identified as COVID-19 positive via RT-PCR test and 3010 negative blood samples were collected from 3010 individuals. For the Ag test, 62 positive and 70 negative samples identified as COVID-19 were collected through RT-PCR testing. Each population was categorized by patient specimen collection dates, the presence of COVID-19 symptoms, pre-existing disease, and age.

**Table 1.** Clinical characteristics of study population.

|  |  | SARS-CoV-2 Ab test |  | SARS-CoV-2 Ag test |  |
| --- | --- | --- | --- | --- | --- |
|  |  | Positive | Negative | Positive | Negative |
| Total |  | 362 | 3010 | 62 | 70 |
| Age range (avg.) |  | 1-88 (59.8) | - | - | - |
| Disease | Y | 207 | - | - | - |
|  | N | 155 | - | - | - |
| Days post symptom | 0-7 | 40 | - | - | - |
|  | 8-14 | 70 | - | - | - |
|  | 15-21 | 61 | - | - | - |
|  | 22+ | 191 | - | - | - |

### Result of SARS-CoV2 Ab test

In order to compare the positive rate of PCL COVID19 Total Ab EIA and PCL COVID19 IgG/IgM Rapid Gold to RT-PCR, the obtained results were first categorized into 4 groups by the length of days after symptom onset: 0-7 days, 8-14 days, 15-21 days, and 22+ days. The positive rate of PCL COVID19 Total Ab EIA diagnostic results for 362 samples, confirmed positive with RT-PCR, was 40% (95% CI 24.86-56.67%), 65.7% (95% CI 53.40-76.65%), 93.4% (95% CI 84.05-98.18%) and 94.2% (95% CI 89.93-97.09%) respectively. For IgG & IgM combined positive rates of PCL COVID19 IgG/IgM Rapid Gold diagnostics for 362 identical samples was 42.5% (95% CI 27.04-59.11%), 64.3% (95% CI 51.93-75.39%), 86.9% (95% CI 75.78-94.16%), and 97.4% (95% CI 94.00-99.14%) respectively. We also measured the positive rate marked with IgG and IgM individually; IgG showed a result of 32.5% (95% CI 18.57-49.13%), 50.0% (95% CI 37.80-62.20%), 83.6% (95% CI 71.91-91.85%) and 97.4% (95% CI 94.00-99.14%), and IgM showed 37.5% (95% CI 22.73-54.20%), 51.4% (95% CI 39.17-63.56%), 77.0% (95% CI 64.5-86.85%), and 65.4% (95% CI 58.24-72.16%) accordingly. This data demonstrated that IgG and IgM showed a similar positive rate until the 14 days period, but we noticed that the positive rate of IgG began to gradually increase from 14 days and onwards (Figure1).

For PCL COVID19 Total Ab EIA, specificity was measured with a total of 3010 individual serum samples. Only 1 out of 3010 negative samples was a false negative, resulting in specificity of 99.97% (95% CI 99.82-100%) (Table 2). For PCL COVID19 IgG/IgM Rapid Gold, out of 3010 negative samples, 2954 samples were diagnosed as negative and the rest of 56 samples came out as false positives, showed a specificity of 98.14% (95% CI 97.59-98.59%) (Table2). Within the 50 COVID-19 positive patient groups, seroconversion has occurred to 16 of them and we decided to analyze the effect of seroconverted samples on positive rate results of PCL COVID19 Total Ab EIA diagnostic kit. This data was also categorized by the length of days after symptom onset of the patient: 0-7 days, 8-14 days, 15-21 days, and 22+ days, and the result showed as 17.4% (95% CI 4.95-38.78%), 51.2% (95% CI 35.13-67.12%), 90.5% (95% CI 69.62-98.83%), and 92.1% (95% CI 84.99-96.52%) respectively (Figure 2). For antibody detection diagnostics, we observed that the positive rate dramatically increases after 14 days from the infection of the virus.

**Table 2.** Results of negative test for each kit.

|  | PCL COVID19 Total Ab EIA | PCL COVID19 IgG/IgM Rapid gold | PCL COVID19 Ag Rapid FIA |
| --- | --- | --- | --- |
| Total sample | 3010 | 3010 | 70 |
| Positive | 1 | 56 | 1 |
| Negative | 3009 | 2954 | 69 |
| Specificity | 99.97% | 98.14% | 98.57% |

**Figure 1.**
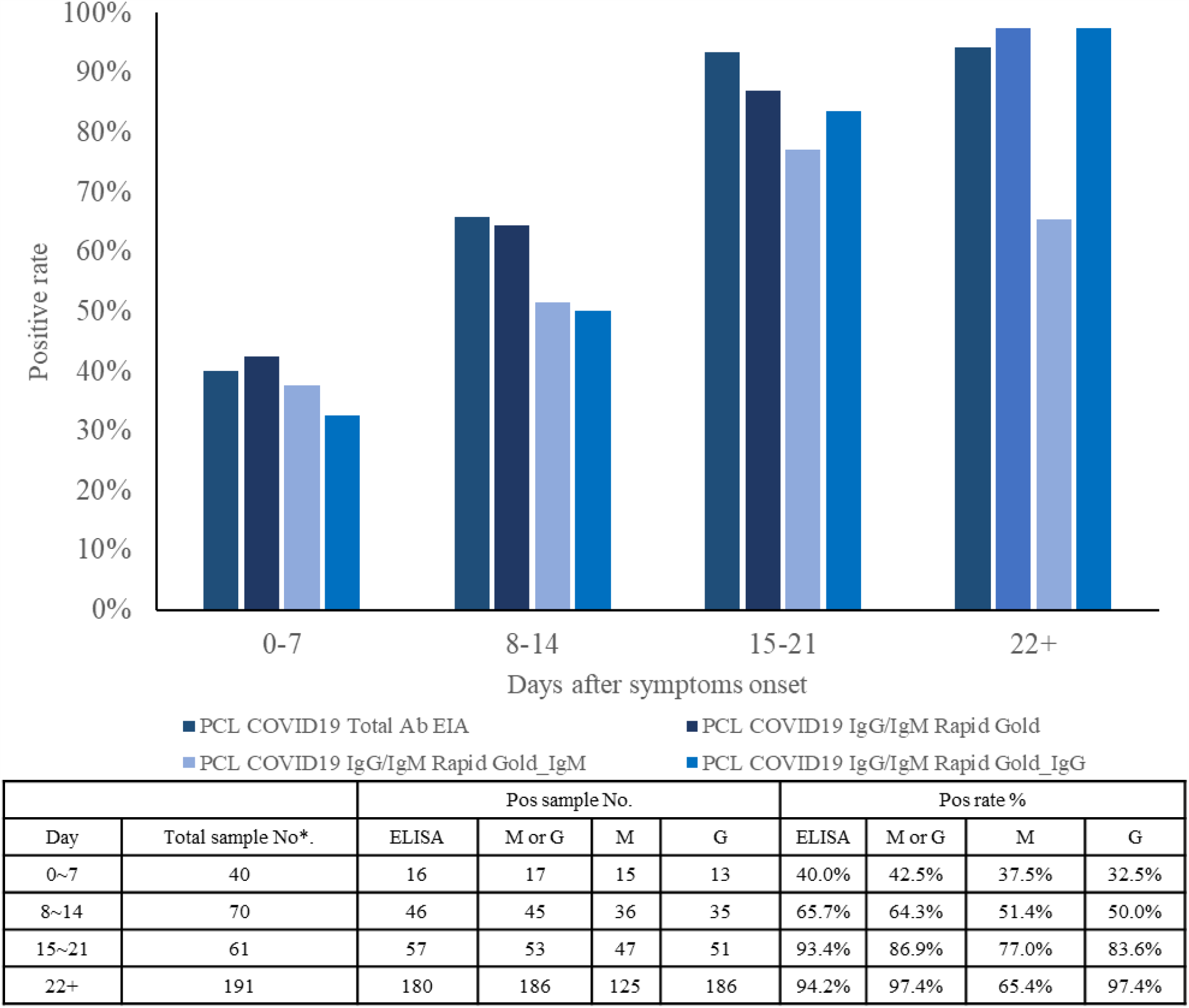
Graph and table of positive rates of total antibody, IgG and IgM and days after symptom onset. Positive rates for the total antibody(measured by PCL COVID19 Total Ab EIA), IgG/IgM(measured by PCL COVID19 IgG / IgM Rapid Gold), were determined in 362 serum samples from 50 patients according to the days after symptom onset.

**Figure 2.**
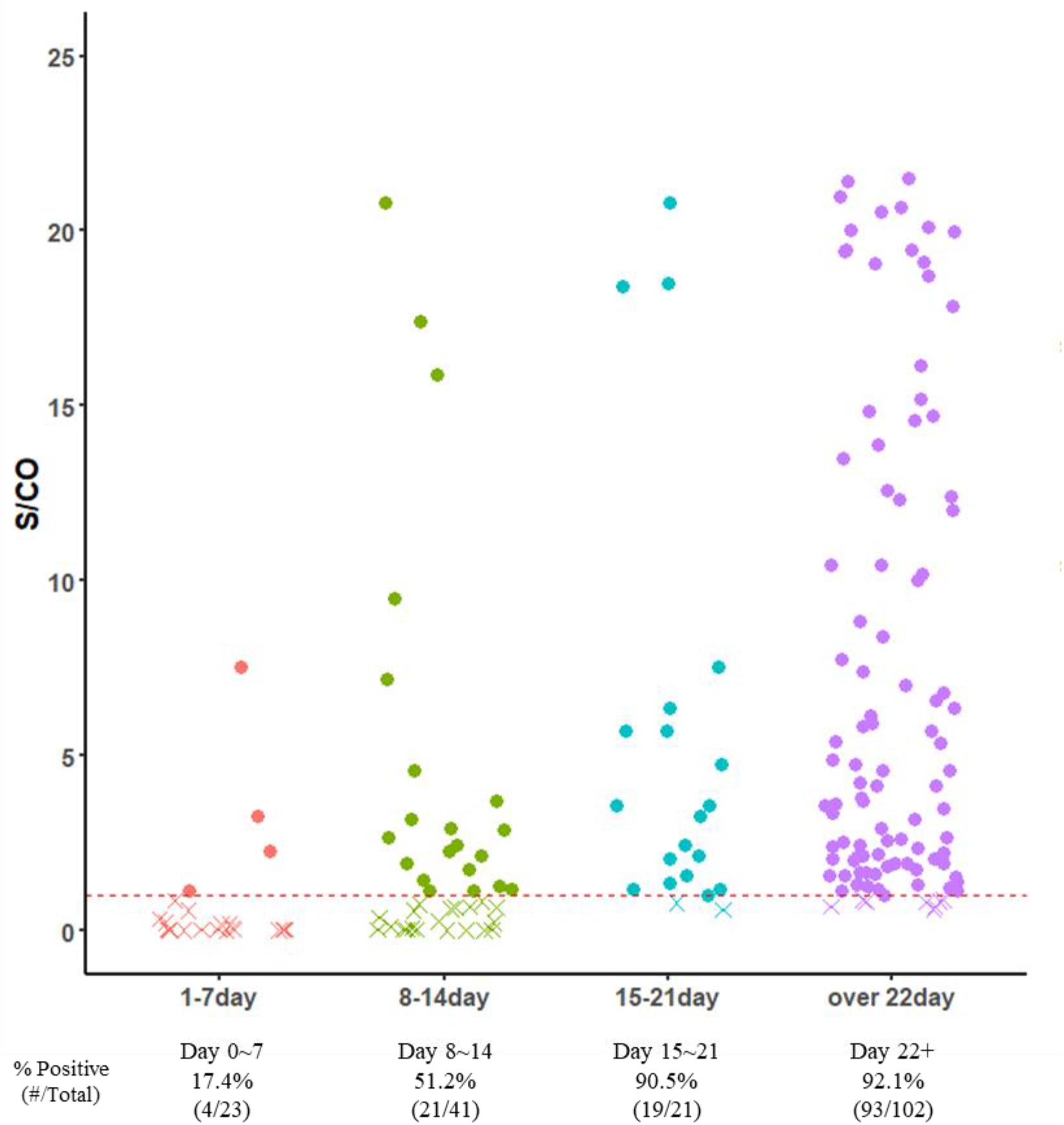
Levels of antibodies against SARS-CoV-2 in patient samples at different times after symptom onset. SARS-CoV-2 antibody levels were measured for various periods in 16 patients with seroconversion, and the positive rate of antibodies measured within each week was plotted on the graph. The X shape is less than S/CO 1, the point is S/CO 1 or more. The red dotted line is S/CO 1(equal to cutoff)

In addition, by comparing the results from patients with the presence or absence of disease (Figure 4B), it was confirmed that the antibody concentration was high in patients with underlying disease.

Taken together, it seems that SARS-CoV-2 antibody should be tested after 14 days of infection, and SARS-CoV-2 antigen should be tested within 14 days of infection.

### Result of SARS-CoV2 Ag test

We used PCL COVID19 Ag Rapid FIA to distinguish the patients who have been infected for less than 14 days. With having all 62 samples already confirmed positive with RT-PCR test (0-12 days after symptom onset), the test result demonstrated that sensitivity was 93.8% (95% CI 82.80-98.69%) for 0-7 days period and 71.4% (95% CI 41.90-91.61%) for 8-12 days period (Figure 3). We can conclude that the positive rate for antigen-based tests decreases as days go by from the initial symptom onset period of a patient. Specificity was measured with a total of 70 RT-PCR confirmed negative samples, and only 1 out of 70 samples was false negative, resulting in specificity of 98.57% (95% CI 92.30-99.96%) (Table 2).

**Figure 3.**
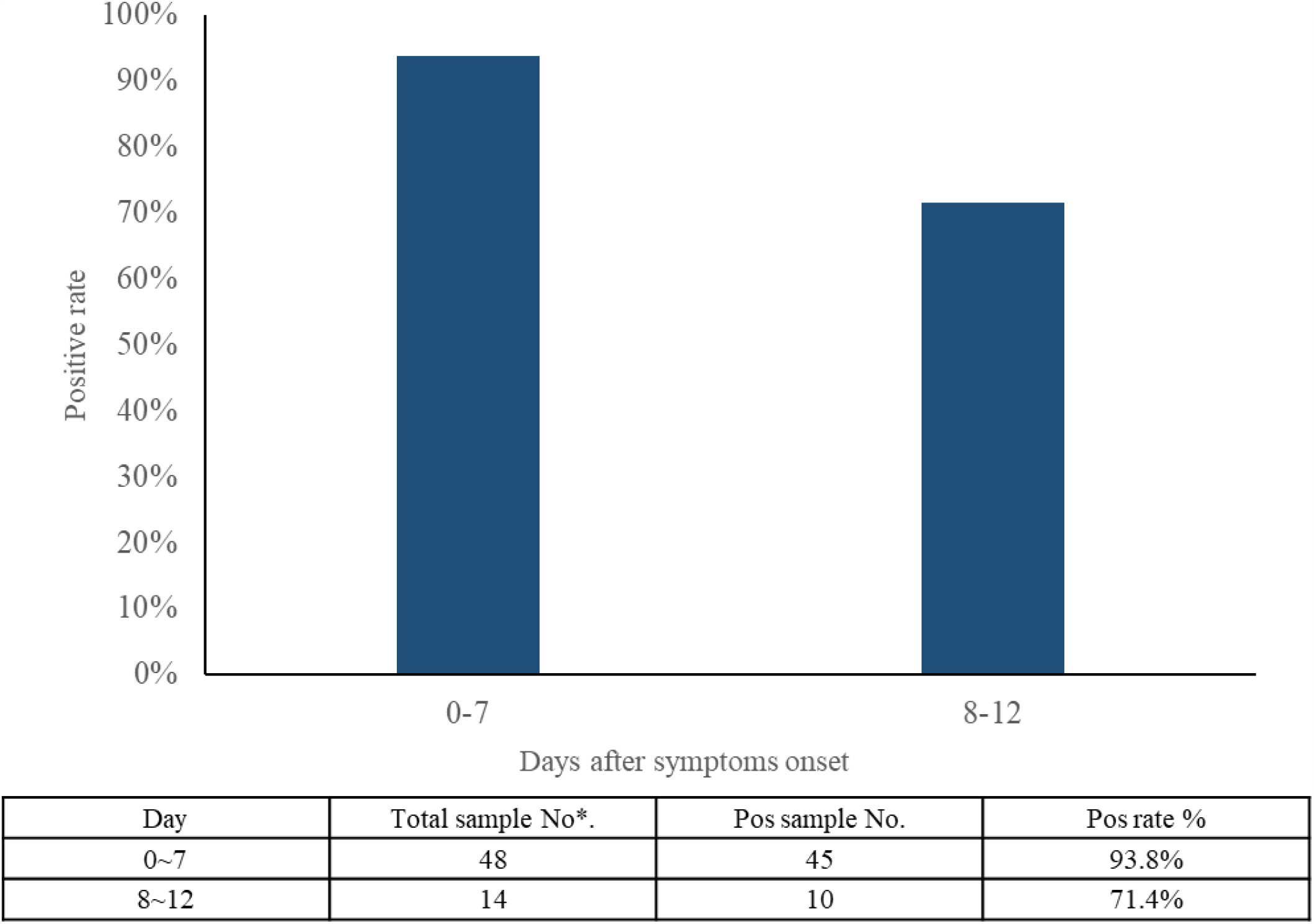
Graphs of positive rates and days after symptom onset of PCL COVID19 Ag Rapid FIA. The positive rate is the result of PCL COVID19 Ag Rapid FIA diagnosis, according to the number of days after onset in 62 RT-PCR positive diagnostic samples.

**Figure 4.**
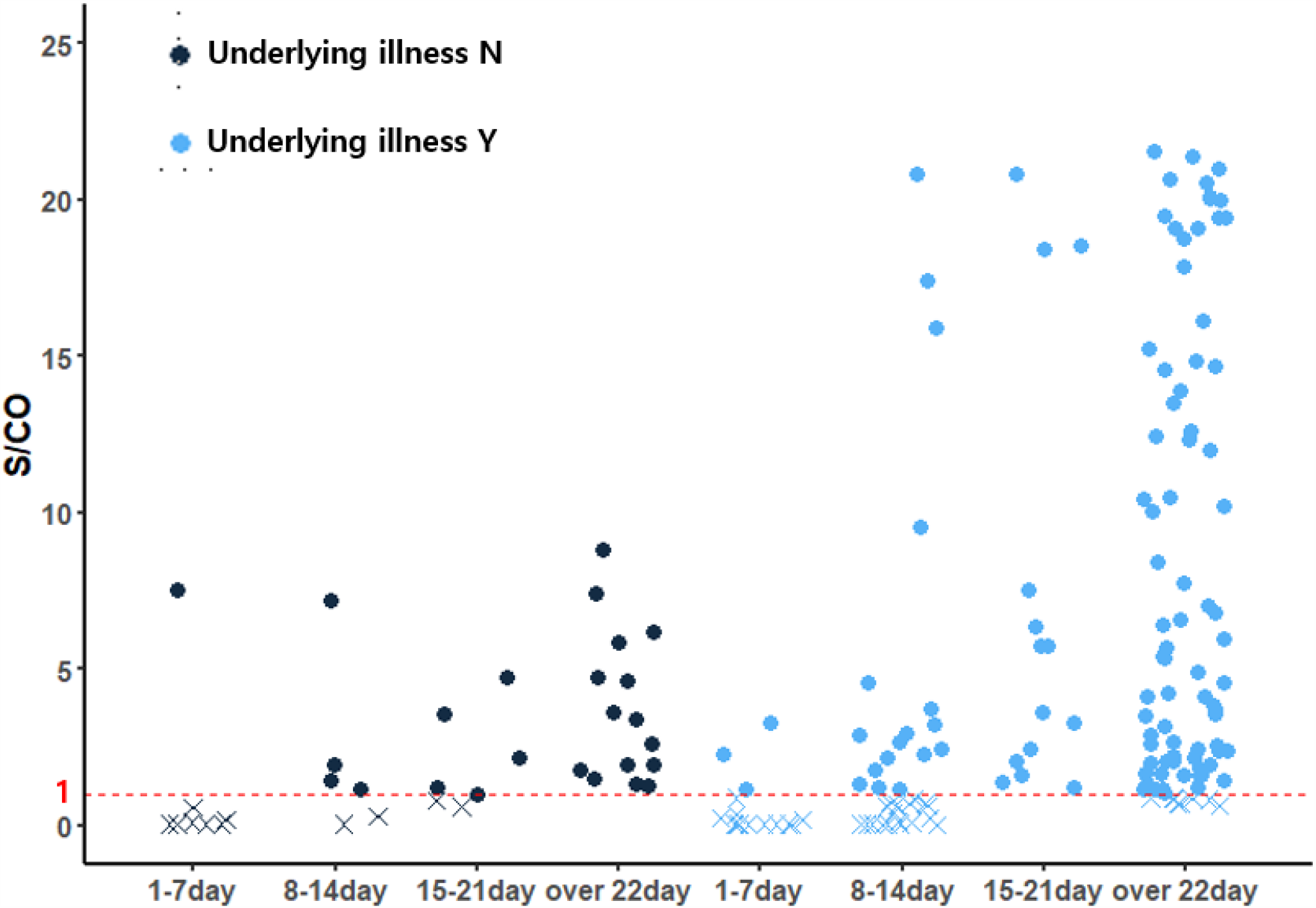
Antibody levels against SARS-CoV-2 according to the clinical characteristics of the study population. Levels of antibodies against SARS-CoV-2 between patients with or without underlying illness at different times after symptom onset. The X shape is less than S/CO 1, the point is S/CO 1 or more. The red dotted line is S/CO 1(equal to cutoff). 16 patients with seroconversion(PCL COVID19 Total Ab EIA result) were picked up and studied.

## Discussion

In Wuhan, Hubei Province, China, coronavirus disease 2019 (COVID-19) was declared a pandemic in March 2020 by the World Health Organization (WHO) with the first detection of extreme acute respiratory syndrome coronavirus 2 (SARS-CoV-2) in December 20191. The WHO announced that Chinese authorities had identified a novel type of coronavirus in January 7 2020^13^. RT-PCR, a nucleic acid amplification test (NAAT), was used as a COVID-19 diagnostic test in many countries around the world, since the new coronavirus was identified4-7. However, the RT-PCR test takes a lot of time to accurately diagnose the infection. To combat the current COVID-19 pandemic, an efficient strategy for diagnosis of asymptomatic infections is required because it is difficult to discern solely with a symptom-based RT-PCR screening. Asymptomatic cases are reported to be 20 to 40% of the total incidence, or even higher, considering the percentage of undetected COVID-19 infections^14, 15^. Although there have been no cases previously with the Middle East Respiratory Syndrome coronavirus (MERS-CoV), current investigations have shown that asymptomatic and mild COVID-19 patients can produce detectable levels of antibodies against SARS-CoV-19. To manage these problems, the need for an alternative analysis that enables a large number of diagnoses in a short time at low cost has emerged. In fact, RT-PCR test does not satisfy the field where antibody results are required because antibody test results cannot be obtained through RT-PCR test. In this study, complementary diagnostic products to RT-PCR will be reviewed for COVID-19 detection. These products are COVID-19 total Ab ELISA: PCL COVID19 Total Ab EIA, COVID-19 spike protein, nucleocapsid protein antibody immunochromatographic rapid test: PCL COVID19 IgG/IgM Rapid Gold, and COVID-19 nucleocapsid protein antigen fluorescent immunoassay rapid test: PCL COVID19 Ag Rapid FIA. All of the above reagents are a dedicated detection reagent for the SARS-Cov 2.

Coronavirus is a single-stranded RNA virus that uses spike protein to enter host cells. Coronavirus is composed of spike protein, membrane protein, envelope protein, and nucleocapsid protein^4, 13^. In general, the diagnosis of COVID-19 infection is made using RT-PCR. While RT-PCR remains an effective microbial diagnostics technique, however, when used alone in the areas of confirming COVID-19, there are some limitations or weaknesses, such as the inability to differentiate virus viability and the development of indeterminate or false-negative results due to low target cell numbers in specimens^16^. To manage these problems, different methods of diagnosis were required for suspected and supervised COVID-19 infections.

From the aforementioned reasons, complementary diagnosis is required in addition to the RT-PCR-based diagnostic method. Among them, COVID-19 Ab Enzyme-Linked Immunosorbent Assay (ELISA), COVID-19 Ab Immunochromatographic rapid test, and COVID-19 Antigen Fluorescent Immunoassay rapid test can be used. ELISA has proven its validity as a diagnostic method for a long time since it was published by Peter Perlmann and Eva Engvall in 1971. The fact that ELSIA can be implemented to a large-scale automatized equipment makes it a suitable complementary diagnosis method regarding the current issue with the COVI-19 pandemic. Immunochromatography was developed with the aim of detecting antigens in blood in the late 1960s and has been validated as a diagnostic method for various infections. The advantages of immunochromatography are its simple test methods, significantly shorter time required to prepare samples and reagents compared to other methods, and inexpensive equipment required for making a diagnosis. Also, it can be applied in various environments both indoor and outdoor settings. Immunochromatography is a suitable complementary diagnosis method for RT-PCR test which has limitations such as operation time, operable environment, and number of samples the test can diagnose at once. Lastly, the COVID-19 Nucleocapsid protein Antigen Fluorescent Immunoassay has gone through major developmental breakthroughs over the years, and has shown prominence in reagent simplification, simple analysis design, and high-sensitivity detection. Fluorescent Immunoassay uses a fluorescent compound that absorbs light or energy of a specific wavelength and emits light or energy at another wavelength. This fluorescent compound offers an advantage of being able to check the results in a short time using a small analyzer as a detection reagent.

PCL COVID19 Total Ab EIA (ELISA) showed the sensitivity of 93.4% from 15 to 21 days, 94.2% from over 22 day, and the specificity of 99.97%. PCL COVID19 IgG/IgM Rapid Gold (ICA) had a sensitivity of 86.9%, 97.4%, and the specificity of 98.14% respectively. PCL COVID19 Ag Rapid FIA sensitivity was 93.8% from 0 to 7 days, 71.4% from 8 to 12 day and specificity was 98.57%. As a result, these diagnostic methods can be used as a complement to the current diagnosis of COVID-19, before 14days of infection, SARS-CoV-2 antigen should be tested and after 14days of infection, SARS-CoV-2 antibody should be tested.

## Data Availability

Due to confidentiality, neither the data nor the source of the data can be made available

## Acknowledgements

This study was supported by the Promising IP Project Support Program (grant number: 20001089) funded by the Ministry of Trade Industry and Energy. And this study was supported by the Clinical Trial Support Program (grant number: HI18C1849) funded by the Ministry of Health and Warfare. Also, this study was supported by the Technological Innovation R&D Program (grant number: HI20C0371010120) funded by the Korea Health Industry Development Institute (KHIDI).

